# Estimating individual risk of catheter-associated urinary tract infections using explainable artificial intelligence on clinical data

**DOI:** 10.1101/2024.03.22.24304712

**Authors:** Herdiantri Sufriyana, Chieh Chen, Hua-Sheng Chiu, Pavel Sumazin, Po-Yu Yang, Jiunn-Horng Kang, Emily Chia-Yu Su

## Abstract

**Background:** Catheter-associated urinary tract infections (CA-UTIs) significantly increase clinical burdens. Identifying patients at high-risk of CA-UTIs is crucial in clinical practice. In this study, we developed and externally validated an explainable, prognostic prediction model of CA-UTIs among hospitalized individuals receiving urinary catheterization.

**Methods:** We applied a retrospective cohort paradigm to select data from a clinical research database covering three hospitals in Taiwan. We developed a prediction model using data from two hospitals and used the third hospital’s data for external validation. We selected predictors by a multivariate regression analysis through applying a Cox proportional-hazards model. Both statistical and computational machine learning algorithms were applied for predictive modeling: (1) ridge regression; (2) decision tree; (3) random forest (RF); (4) extreme gradient boosting; and (5) deep-insight visible neural network. We evaluated the calibration, clinical utility, and discrimination ability to choose the best model by the validation set. The Shapley additive explanation was used to assess the explainability of the best model.

**Results:** We included 122,417 instances from 20-to-75-year-old subjects with multiple visits (*n*=26,401) and multiple orders of urine catheterization per visit (*n*=35,230). Fourteen predictors were selected from 20 candidate variables. The best prediction model was the RF for predicting CA-UTIs within 6 days. It detected 97.63% (95% confidence interval [CI]: 97.57%, 97.69%) CA-UTI positive, and 97.36% (95% CI: 97.29%, 97.42%) of individuals that were predicted to be CA-UTI negative were true negatives. Among those predicted to be CA-UTI positives, we expected 22.85% (95% CI: 22.79%, 22.92%) of them to truly be high-risk individuals. We also provide a web-based application and a paper-based nomogram for using the best model.

**Conclusions:** Our prediction model was clinically accurate by detecting most CA-UTI positive cases, while most predicted negative individuals were correctly ruled out. However, future studies are needed to prospectively evaluate the implementation, validity, and reliability of this prediction model among users of the web application and nomogram, and the model’s impacts on patient outcomes.

## Introduction

Catheter-associated urinary tract infections (CA-UTIs) are among the most common healthcare-associated infections. UTIs contribute 12.19% to health care-associated infections, of which 70% of them are CA-UTIs [1]. Such infections occur when bacteria ascend the urinary tract. Biofilm formation on the catheter surface can harbor micro-organisms, and cause them to be resistant to antibiotic treatment. CA-UTIs significantly increase healthcare costs due to prolonging hospital stays, and requiring additional diagnostic tests and relevant treatments. Overuse of antibiotics to treat CA-UTIs can further lead to increased antimicrobial resistance. CA-UTIs can cause serious complications like sepsis and increased mortality, especially in vulnerable populations such as the elderly or those with comorbid conditions. Preventing CA-UTIs involves minimizing the use of catheters, using aseptic techniques for catheter insertion and maintenance, and removing catheters as soon as clinically feasible. Education and training of healthcare workers in catheter management are crucial.

A high-risk subpopulation of this condition is patients in intensive care units (ICUs) [2]. CA-UTIs increased the length of stay in ICUs by 1.59 days and the mortality risk in ICUs by 15% (95% confidence interval [CI]: 3%, 28%). However, 72% of CA-UTIs occur in patients outside of ICUs. Therefore, preventing CA-UTIs in all hospitalized settings is crucial. Ongoing research is essential to understanding the evolving epidemiology of CA-UTIs and developing new prevention and treatment strategies. Previous studies used multidisciplinary teams, daily catheter reviews, protocol and procedural changes, electronic order sets, catheter insertion and removal reminders, audits and feedback, and education to prevent CA-UTIs [3]. However, these interventions are systemic in nature, but lack personalization. Therefore, identifying patients at high-risk of CA-UTIs in all hospitalized settings is essential, particularly for a specific preventive strategy of CA-UTIs at the individual level, e.g., antimicrobial catheter usage.

Only a few prediction models have been developed on relevant issues. Two models for predicting CA-UTIs were developed among patients in ICU settings for neurologic cases only, applying statistical and computational machine learning (ML) algorithms. The first ML model achieved an area under curve (AUC) of the receiver operating characteristics (ROC) of 0.888 (95% CI: 0.772, 1.000) by a validation set [4]. Meanwhile, the second ML model achieved an AUC-ROC of 0.920 (95% CI: 0.786, 0.918) by a training set [5]. Unfortunately, no previous study developed and validated a prediction model of CA-UTIs among all hospitalized population.

This risk prediction is expected to pave the way to a more-effective approach for combating CA-UTIs. Furthermore, the model’s explainability may guide reliable decision-making by reducing critical risk factors. In this study, we aimed to develop and externally validate an explainable, prognostic prediction model of CA-UTIs among hospitalized individuals receiving urinary catheterization.

## Methods

### Study design

We applied a retrospective cohort paradigm to select data from the Taipei Medical University (TMU) Clinical Research Database (TMUCRD) (access approval no.: A202206008). This integrated research database consists of clinical data from deidentified and curated electronic medical records (EMRs) in three hospitals: (1) Shuang Ho hospital; (2) TMU hospital, and (3) Taipei Municipal Wanfang hospital. The database period is up to 23 years (1998 to 2021) and covers 4.1 million patients across Taiwan. The Taipei Medical University Joint Institutional Review Board gave ethical approval for this work (TMU-JIRB no.: N202209030).

Data of a subject were selected for our cohort if they met the following criteria: (1) the subject’s age ranged from 20 to 75 years, and (2) urinary catheterization was prescribed for the individual. We included all data from the time of catheterization initiation to 48 h post-removal, a period henceforth referred to as an ’episode’. In instances where a catheterization episode concluded within one day or less before commencement of a subsequent episode for the same subject during a hospital visit, these episodes were amalgamated into a single episode. This process was recursively applied until the interval between episodes exceeded one day or no further catheterization occurred. The observational unit was each day within a catheterization episode. A case of a CA-UTI was identified if (1) a diagnosis of UTI and/or an order for a urinary bacterial culture was recorded, and (2) there was no concurrent diagnosis of pneumonia. Both criteria had to be met within the period of catheterization during hospitalization.

Our prediction model was developed to aid decision-making in preventing CA-UTIs. To achieve this objective, we aimed to establish an optimal risk probability threshold. Our approach in determining this threshold prioritized minimizing the rate of false positives (targeting an approximately 10% false positive rate, FPR), as we considered false positives more detrimental than false negatives in the context of this study due to potential iatrogenic harm. While we anticipated this threshold to yield an approximate specificity of 90%, our foremost concern was to ensure significant clinical utility. This utility was assessed by evaluating the net benefit, which was higher than scenarios where all patients were universally treated or categorized as positive (treat all) or not treated or as negative (treat none). In addition to this, we also focused on evaluating the model’s calibration and discrimination capabilities to ensure that the model provided confident predictions and accommodated a broad range of thresholds.

### Predictive modeling

We pre-determined 20 candidate predictors based on a literature review, which were hypothetically considered to be the candidate predictors of outcomes including: (1) age group (≤50/>50 to 65/>65 years old) [6]; (2) ICU stay ≥10 days (no/yes) [2]; (3) duration of catheterization ≥6 days (no/yes) [6]; (4) hospital stay ≥4 days (no/yes) [7, 8]; (5) previous catheterization (*n*) [8]; (6) sex (female/male) [8–11]; (7) type-2 diabetes mellitus (T2DMl no/yes) [8, 10, 12]; (8) immobilization (no/yes) [12]; (9) indication of catheterization (surgical/medical) [10, 12]; (10) drainage system [10, 11]; (11) DM-related factors (no/yes) [13]; (12) poor nutrition (no/yes) [9, 10]; (13) fecal incontinence medication (no/yes) [9]; (14) impaired immune function (no/yes) [9]; (15) serum creatinine >2 mg/dL (no/yes) [9, 10]; (16) sodium-glucose co-transporter (SGLT)-2 inhibitors (no/yes) [14]; (17) urological issues (no/yes) [15, 16]; (18) previous UTI diagnosis >2 times per six months or >3 times per year (no/yes) [17]; (19) neurological issues (no/yes) [18]; and (20) antibiotic drugs last seven days (no/yes) [10]. Data availability of these candidate predictors was identified after partitioning our data. In addition, the duration of catheterization as a candidate predictor was defined as days from the start of urinary catheterization to the day of prediction.

To conduct data partition, we used data from the TMU and Shuang Ho hospitals for the development set and those from the Taipei Municipal Wanfang hospital for the test set.

Subsequently, we randomly split the development set into training and validation sets by an80:20 ratio. The downstream analysis only used the training set, except when internally (i.e., by the validation set) and externally (i.e., by the test set) validating a prediction model.

Before predictor selection, we unselected a candidate predictor with (1) unavailable data,(2) perfect separation problem, and (2) missing not at random (MNAR). A categorical candidate predictor was considered to perfectly separate the outcome if there were no samples in any combinations of categories between the candidate predictor and the outcome. With the exception of data generation that allowed perfect separation (e.g., no CA-UTI sample among patients without urine catheterization), it was considered a sampling error (simply occurring by chance). A predictive modeling algorithm may falsely choose such a candidate predictor as an important predictor. Meanwhile, a candidate predictor was considered MNAR if there was a significant association between its missing value (not missing/missing) and the outcome (negative/positive/missing). For a numerical candidate predictor, we also identified outliers.

However, we removed no samples with outliers. Instead, we assigned missing values to the outliers, assuming that these might be a random error when the data were input during patient care. Subsequently, we reconsidered if a candidate predictor was MNAR.

Furthermore, if a candidate predictor was unselected and had either >1 indicators or a numerical indicator, we substituted it with any of its indicators. They were also unselected if they fulfilled the above criteria for candidate predictor selection. Eventually, multiple imputations by a chained equation were applied to missing values of the selected candidate predictors and the outcome and its onset. An outcome onset was determined by how many days a patient was diagnosed with a CA-UTI after the start of a urinary catheterization. Although we included the outcome and its onset for imputing candidate predictors, we did not use the imputed values of the outcome or its onset. Only data with a non-missing outcome were used. We included missing values when computing the outcome probabilities for inverse probability weighting in the predictive modeling. In addition, the numerical candidate predictors were also standardized by their respective average and standard deviation (SD), which were computed without outliers. A numerical predictor standardization utilized the training set’s corresponding average and SD in the validation and test sets.

For predictor selection in our study, we employed Cox proportional-hazards modeling for both the univariate and multivariate regression analyses. Initially, a univariate regression analysis was conducted to ascertain associations between each candidate predictor and the outcome of interest. For the multivariate regression analysis, we assessed interrelationships among selected candidate predictors through either a linear or logistic regression analysis. A variable was designated as a covariate for a particular candidate predictor if it demonstrated an association with both the candidate predictor and the outcome in the multivariate analysis. Given the hypothesized causal direction from the candidate predictor to the outcome, the identified covariates were categorized as either confounders or mediators based on the principles of structural causal modeling. Consequently, in the multivariate regression, a candidate predictor’s hazard ratio (HR) might represent either its direct effect or total effect, excluding the effect of covariates included in this study.

These predictor selection procedures were also applied across various combinations of outcome onset spans and ranges. For instance, a span of 3 days and a range of 15 days imply assessing the outcome every three days for up to 15 days. We considered 1, 3, and 7 days for outcome evaluations, with corresponding outcome onset ranges of 5 to 90 days. The minimum range (5 days) ensured at least five groups of outcome onset for each span, which was deemed adequate for Cox proportional-hazards modeling. The maximum range (90 days) was set as the acceptable upper limit of follow-up to identify positive CA-UTI cases. The selection of a combination of span and onset was based on the minimum sample size criterion among outcome onset groups for each span, with preference given to the largest minimum sample size. The combination with the longest range was chosen in case of a tie.

Given the extensive application of predictor selection across various combinations of outcome onsets and spans, the possibility of a multiple-testing effect was acknowledged. To address this, we corrected *p*-values of association tests by the Benjamini-Hochberg method, thereby controlling the false discovery rate. A candidate predictor was selected only if it showed a significant association with the outcome after adjusting for covariates (if any), signified by a corrected *p*-value of ≤0.05. We only utilized predictors selected via the multivariate regression analysis to develop the prediction models. The models were tailored to follow-up within the selected range and span. For example, with a chosen range of 15 days and a span of 3 days, four models were developed to predict the outcome without and at specific onset intervals (days 1 to 3, 6, 9, and 12). In cases where a categorical predictor had more than two categories, it was transformed into multiple predictors, excluding the reference category.

To construct the prediction models, we applied five ML algorithms to develop a prediction model: ridge regression (RR), decision tree (DT), random forest (RF), extreme gradient boosting (XGBoost), and deep-insight visible neural network (DI-VNN). These algorithms were selected for their diverse approaches and various handling of data characteristics, as well as their hyperparameters (Table S1). RR, a statistical ML algorithm, is notable for its linear approach and L2-norm regularization. In contrast, DT, RF, XGBoost, and DI-VNN are computational algorithms that primarily address non-linearity. DT operates by stepwise classification based on predictor importance, whereas RF aggregates multiple DTs for outcome classification. XGBoost sequentially refines DTs to minimize misclassification rates, and DI-VNN, a deep learning algorithm, includes inter-predictor relationships in its predictions, a feature not commonly found in other ML approaches. The predictive performance of models developed with these algorithms varied depending on the data characteristics. Therefore, we conducted a robust statistical analysis to compare these models and selected the most effective for predictive modeling in our study.

### Statistical analysis

We assessed the respective calibration, clinical utility, and discrimination ability of the models to robustly evaluate them. We screened the well-calibrated models that demonstrated a higher net benefit relative to the treat-all and treat-none scenarios (see Study design) and a better discriminative ability than a random guess. We chose a prediction model with the best discriminative ability among the models that passed the screening. The explainability of the best model was also evaluated. As mentioned above, all evaluation metrics were assessed from the validation set to choose the best model. We also showed the evaluation results based on the test set for the best model to ensure the robustness of its predictive performance; hence, it allowed the model to be deployed in real-world settings. We inferred the interval estimate (i.e., the 95% CI) for each evaluation metric by 30-time bootstrapping. We shared the programming codes for all analyses in this study (https://github.com/herdiantrisufriyana/colab_uti), including predictive modeling.

To assess the model calibration, we binned the predicted probability from 0 to 1 by 0.1 and inferred the point and interval estimates of the true probabilities for each bin. We fitted a regression line over the point estimates on the calibration plot for descriptive purposes. With this regression, a prediction model had a better calibration if the intercept and slope interval estimates respectively included 0 and 1, or their point estimates approached those numbers. The intercept describes if the predicted probabilities under- or over-estimated the true probabilities, implying whether a prediction model covers all of the true predictors. Meanwhile, the slope describes if the increment of the predicted probabilities reflects that of the true probabilities, implying if a prediction model correctly uses information from the available predictors to classify an outcome.

In addition, we also computed the Brier score, i.e., the root-mean-square error between the predicted probabilities and the true values of either positive (=1) or negative CA-UTIs (=0). The best calibrated model was considered one with the lowest Brier score.

Nevertheless, the calibration plot indicated the optimal range of thresholds by which a higher predicted probability classified a sample as CA-UTI positive. This plot also characterized a prediction model in terms of how to use it in clinical settings. The optimal range was considered when assessing a prediction model’s clinical utility and discrimination ability.

Clinical utility was evaluated by the net benefit, which was computed for each of the predicted probabilities from 0 to 1 by 0.01 increments. Within the optimal range of thresholds in terms of calibration, a prediction model is expected to demonstrate net benefits higher than the reference, i.e., the corresponding net benefits using either a treat-all or treat-none scenario. We also identified the optimal range of thresholds with considerably higher net benefits than the reference, and then re-evaluated the calibration within that optimal range.

Before assessing the discrimination ability using a specific threshold, we compared the discrimination abilities among the models by the AUC-ROC. A prediction model’s discrimination ability was considered significantly better than a random guess if the AUC-ROC interval was >0.5. Among the models that passed this criterion, the model with the best discriminative ability had the highest AUC-ROC.

After choosing the best model, we assessed the discrimination ability using a specific threshold for the best model. Using the validation set, we identified which threshold resulted in ∼90% specificity or achieved the expected clinical utility. Four metrics were computed from prediction results made by the chosen threshold using the test set: (1) sensitivity, (2) specificity, (3) positive predictive value (PPV), and (4) negative predictive value (NPV).

We assessed the explainability of the models with the Shapley additive explanation (SHAP) value. We computed these values and interpreted their meaning at both the population and individual levels by beeswarm and waterfall plots. Both estimated each predictor’s impact on the model’s output. At the individual level, we selected four groups of samples that represented true positives, false negatives, false positives, and true negatives. We selected a sample from each distinguished cluster of samples for each group. To determine the clusters, we reduced predictors into two principal components (PCs) with the highest proportions of variance explained. Subsequently, we applied *k*-mean clustering by the PCs with *k* centers such that all visible clusters were separated by classifying one or more clusters. A sample closest to each centroid was taken for each of the groups. Eventually, we expect to learn how to use and improve the best model in the future according to model’s explainability.

We also provided a web application and a nomogram to use the best model. For the latter, we followed a protocol for creating an algorithm-agnostic nomogram [19]. This nomogram allows a paper-based prediction, increasing the accessibility of the best prediction model, including one developed by a complex ML algorithm.

## Results

### Baseline characteristics

We identified 20-to-75-year-old subjects (*n*=18,164) with one or more visits (*n*=26,401) to the three hospitals and with one or more orders of urinary catheterization per visit (*n*=35,230) (Figure 1A). This selection resulted in 122,417 instances for developing and validating our prediction models, i.e., days of prediction within all catheterization episodes. Subjects visiting Taipei Municipal Wanfang hospital constituted 20.85% of the instances, i.e., the test set, while the remaining subjects were from Shuang Ho and TMU hospitals, i.e., the development set. The prevalence of CA-UTIs in the test set was considered higher than that in the development set (*n*=13,432/101,294 [19.29%] vs. *n*=4074/21,123 [13.26%]), which was a good challenge to test the robustness of the best model. Eventually, we randomly split the development set into the training and validation sets (*n*=10,676/81,010 [79.98%] vs. *n*=2756/17,497 [20.02%]).

**Figure 1.**
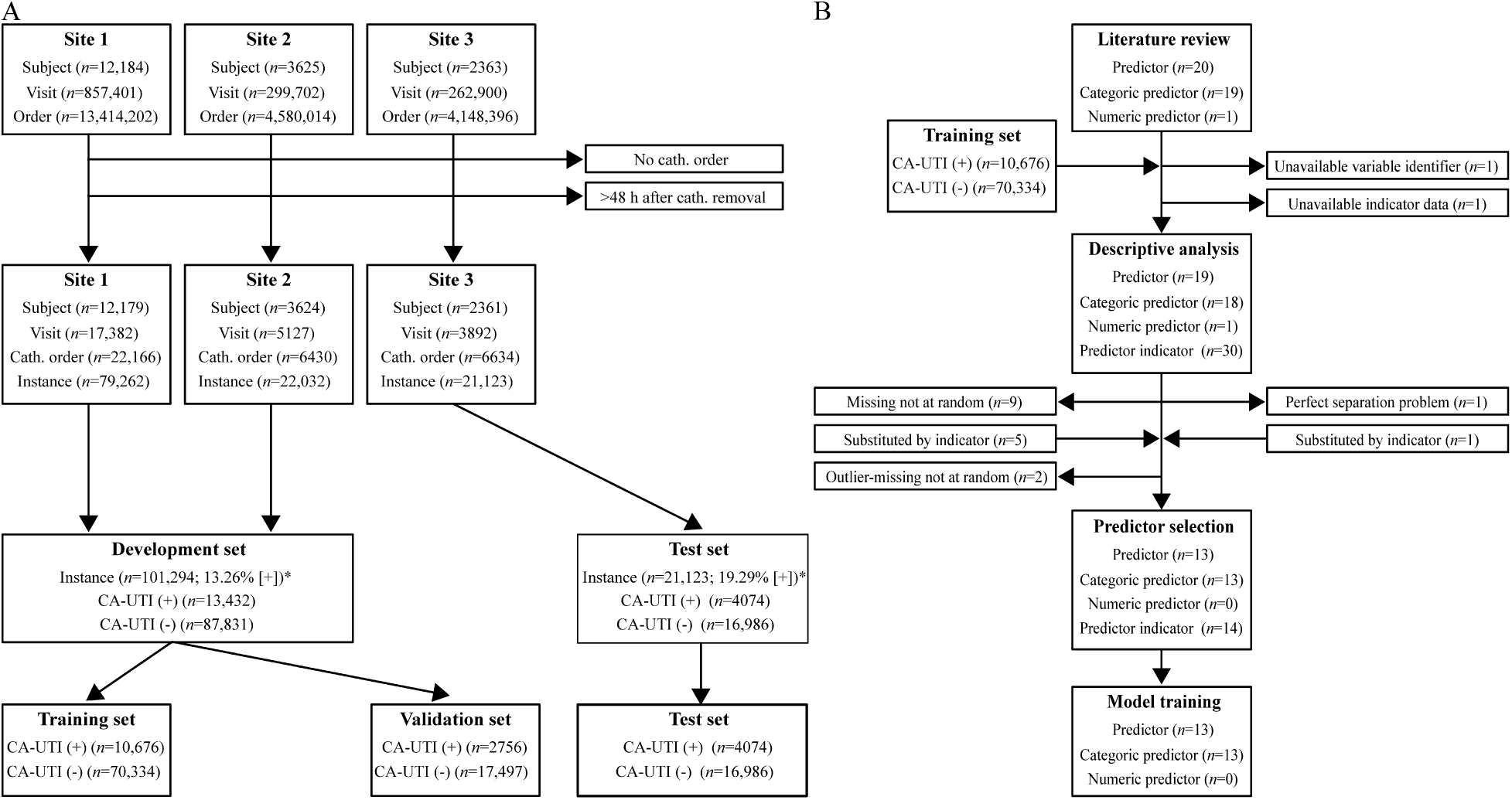
Data selection: (A) observations; and (B) predictors. Site 1, 2, and 3 are respectively the Shuang Ho hospital, the TMU hospital, and the Taipei Municipal Wanfang hospital. *, including missing values; Cath., catheterization; CA-UTI, catheter-associated urinary tract infection.

We identified 30 indicators for 19 candidate predictors in the training set (Table S2; Figure 1B). We unselected the duration of catheterization ≥6 days since it perfectly separated the outcome. While we had substituted this candidate predictor by its indicator, i.e., duration of catheterization in days, we also unselected this candidate predictor since it was MNAR after assigning its outliers as missing. The nine other candidate predictors were also MNAR, some of which were substituted by five indicators, i.e.: (1) T2DM (substituted by T2DM diagnosis only, excluding DM medication); (2) indication of catheterization (no substitution); (3) DM-related factors (substituted by either hypertension or DM with nephropathy); (4) poor nutrition (the remaining indicators were also MNAR before or after assigning their outliers as missing); (5) fecal incontinence medication (its indicators were also MNAR); (6) impaired immune function (substituted by either human immunodeficiency virus [HIV] or malignant neoplasms); (7) serum creatinine >2 mg/dL (its numerical indicator was also MNAR after assigning its outliers as missing); (8) SGLT-2 inhibitors (no substitution); and (9) antibiotic drugs last seven days (no substitution). The previous catheterization was unselected since it was MNAR after assigning its outliers as missing. Eventually, thirteen candidate predictors passed the preliminary check to be included in predictor selection.

In the training set (Table 1), most CA-UTI positives were characterized by the >50-to-65-year-old age group, ICU stay <10 days, hospital stay ≥4 days, female, or negative for other selected candidate predictors. Similar characteristics were found among CA-UTI negatives, except most were characterized by the ≤50-year-old age group or hospital stay <4 days. Unlike the validation set with similar characteristics to those in the training set, CA-UTI positives and negatives in the test set were characterized by the >50-to-65-year old age group or hospital stay ≥4 days (Table S3).

**Table 1.**
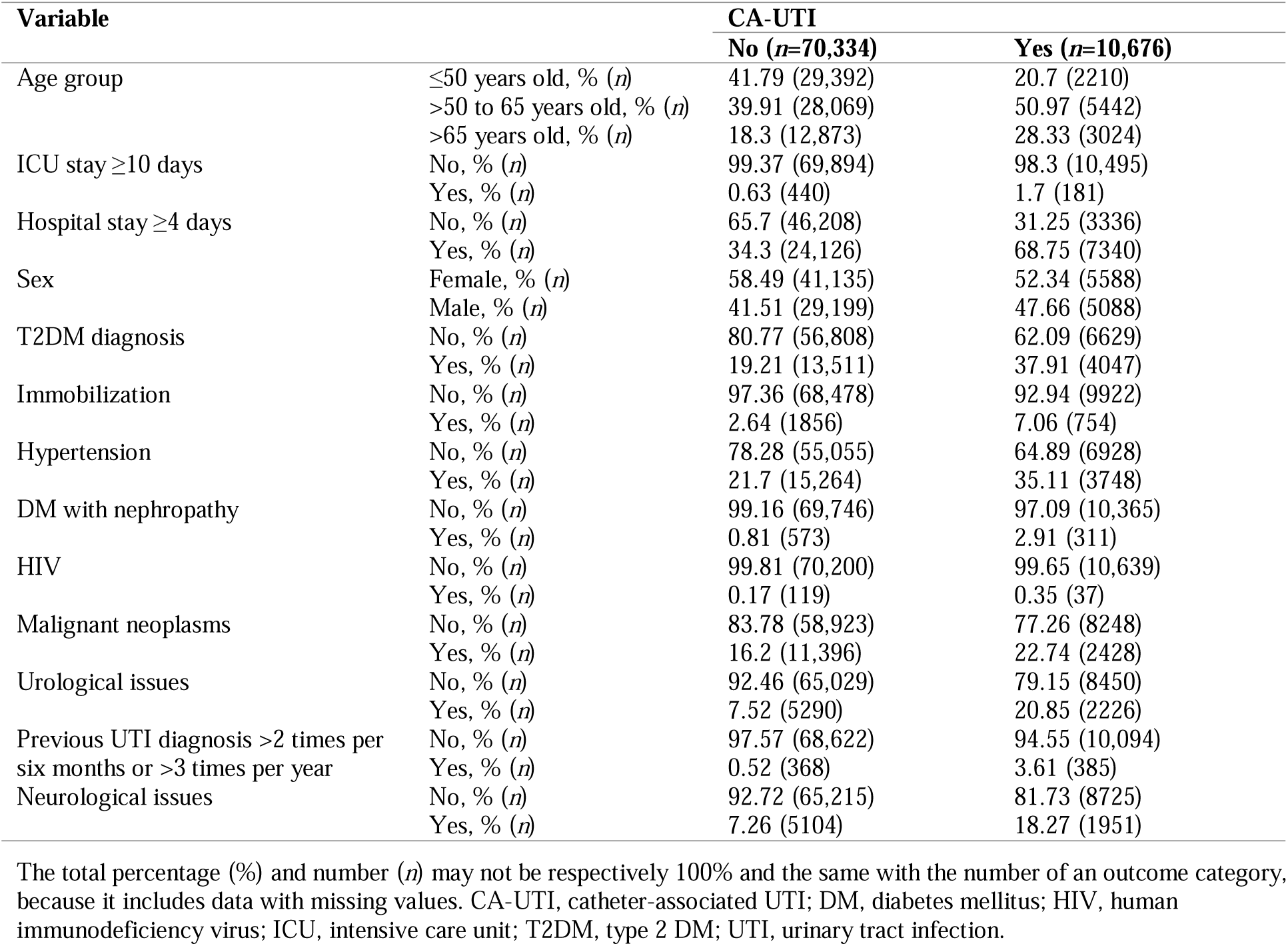
Baseline characteristics of training set.

| Variable |  | CA-UTI |  |
| --- | --- | --- | --- |
|  |  | No (n=70,334) | Yes (n=10,676) |
| Age group | ≤50 years old, % (n) | 41.79 (29,392) | 20.7 (2210) |
|  | >50 to 65 years old, % (n) | 39.91 (28,069) | 50.97 (5442) |
|  | >65 years old, % (n) | 18.3 (12,873) | 28.33 (3024) |
| ICU stay ≥10 days | No, % (n) | 99.37 (69,894) | 98.3 (10,495) |
|  | Yes, % (n) | 0.63 (440) | 1.7 (181) |
| Hospital stay ≥4 days | No, % (n) | 65.7 (46,208) | 31.25 (3336) |
|  | Yes, % (n) | 34.3 (24,126) | 68.75 (7340) |
| Sex | Female, % (n) | 58.49 (41,135) | 52.34 (5588) |
|  | Male, % (n) | 41.51 (29,199) | 47.66 (5088) |
| T2DM diagnosis | No, % (n) | 80.77 (56,808) | 62.09 (6629) |
|  | Yes, % (n) | 19.21 (13,511) | 37.91 (4047) |
| Immobilization | No, % (n) | 97.36 (68,478) | 92.94 (9922) |
|  | Yes, % (n) | 2.64 (1856) | 7.06 (754) |
| Hypertension | No, % (n) | 78.28 (55,055) | 64.89 (6928) |
|  | Yes, % (n) | 21.7 (15,264) | 35.11 (3748) |
| DM with nephropathy | No, % (n) | 99.16 (69,746) | 97.09 (10,365) |
|  | Yes, % (n) | 0.81 (573) | 2.91 (311) |
| HIV | No, % (n) | 99.81 (70,200) | 99.65 (10,639) |
|  | Yes, % (n) | 0.17 (119) | 0.35 (37) |
| Malignant neoplasms | No, % (n) | 83.78 (58,923) | 77.26 (8248) |
|  | Yes, % (n) | 16.2 (11,396) | 22.74 (2428) |
| Urological issues | No, % (n) | 92.46 (65,029) | 79.15 (8450) |
|  | Yes, % (n) | 7.52 (5290) | 20.85 (2226) |
| Previous UTI diagnosis >2 times per six months or >3 times per year | No, % (n) | 97.57 (68,622) | 94.55 (10,094) |
|  | Yes, % (n) | 0.52 (368) | 3.61 (385) |
| Neurological issues | No, % (n) | 92.72 (65,215) | 81.73 (8725) |
|  | Yes, % (n) | 7.26 (5104) | 18.27 (1951) |

The total percentage (%) and number (n) may not be respectively 100% and the same with the number of an outcome category, because it includes data with missing values. CA-UTI, catheter-associated UTI; DM, diabetes mellitus; HIV, human immunodeficiency virus; ICU, intensive care unit; T2DM, type 2 DM; UTI, urinary tract infection.

### Predictive performance and explainability

We identified an outcome onset span and range of 3 and 15 days, of which the minimum sample size was the largest among other combinations (Figure S1A). All of the selected candidate predictors were significantly associated with CA-UTIs in the univariate regression analyses (Table 2; Figures S1B to S1M). For each candidate predictor, we identified associations with other candidate predictors (Table S4). After adjustment with other predictors as covariates, the effect sizes of most candidate predictors were significantly smaller on the outcome, except ICU stay ≥10 days, HIV, sex male, hypertension, and malignant neoplasms. Nevertheless, by these multivariate regression analyses, we identified significant associations between each of the selected candidate predictors and CA-UTIs.

**Table 2.** Associations between each of the selected predictors and the outcome.

| Candidate predictor | Association with CA-UTIs by HR (95% CI; *p*-value*) |  |
| --- | --- | --- |
|  | Unadjusted | Adjusted† |
| Age group >50 to 65 years old | 1.981 (1.874, 2.094; <0.001) | 1.490 (1.406, 1.579; <0.001) |
| Age group >65 years old | 2.411 (2.269, 2.563; <0.001) | 1.590 (1.487, 1.700; <0.001) |
| ICU stay $\geq 10$ days | 1.950 (1.656, 2.295; <0.001) | 1.896 (1.611, 2.232; <0.001) |
| Hospital stay $\geq 4$ days | 2.861 (2.734, 2.995; <0.001) | 2.324 (2.216, 2.437; <0.001) |
| Sex male | 1.089 (1.044, 1.136; <0.001) | 0.844 (0.808, 0.881; <0.001) |
| T2DM diagnosis | 1.996 (1.911, 2.084; <0.001) | 1.397 (1.332, 1.466; <0.001) |
| Immobilization | 2.242 (2.069, 2.429; <0.001) | 1.308 (1.204, 1.422; <0.001) |
| Hypertension | 1.610 (1.541, 1.683; <0.001) | 0.942 (0.895, 0.992; 0.023) |
| DM with nephropathy | 2.844 (2.516, 3.214; <0.001) | 1.522 (1.337, 1.733; <0.001) |
| HIV | 2.066 (1.475, 2.893; <0.001) | 2.292 (1.634, 3.215; <0.001) |
| Malignant neoplasms | 1.126 (1.069, 1.185; <0.001) | 0.828 (0.786, 0.873; <0.001) |
| Urological issues | 2.830 (2.689, 2.978; <0.001) | 2.154 (2.039, 2.275; <0.001) |
| Previous UTI diagnosis >2 times per six months or >3 times per year | 4.097 (3.668, 4.577; <0.001) | 2.115 (1.882, 2.377; <0.001) |
| Neurological issues | 2.244 (2.126, 2.368; <0.001) | 1.414 (1.331, 1.501; <0.001) |

\*, after correcting for the multiple testing effect by the Benjamini-Hochberg method; †, covariates for the adjustment are listed in Table S4; CA-UTI, catheter-associated UTI; CI, confidence interval; DM, diabetes mellitus; HR, hazard ratio; HIV, human immunodeficiency virus; ICU, intensive care unit; T2DM, type 2 DM; UTI, urinary tract infection.

We chose RF to predict an outcome with a specific onset from day 1 to 6. By calibration (Figure 2A; Table S5; Figure S2), this model was the only well-calibrated one (intercept -0.193 [95% CI: -0.412, 0.027]; slope 0.844 [95% CI: -0.490, 1.198]; Brier score 0.453 [95% CI: 0.452, 0.453]). The decision curve analysis also demonstrated the optimal range of thresholds for which net benefits were still higher than the references, i.e., maximum ∼0.16 (Figure S3). Such an optimal range was only demonstrated by the RF for predicting an outcome without and with a specific onset from day 1 to 6, 9, and 12. All the prediction models achieved AUCs-ROC of >0.5 (Table S5; Figure S4), among which the RF, XGBoost, and DT did not significantly differ as the most discriminative models for predicting an outcome within 6, 9, and 12 days, or with no specific onset. Hence, we chose the RF for predicting an outcome within six days as the best model according to the calibration, clinical utility, and discrimination ability.

**Figure 2.**
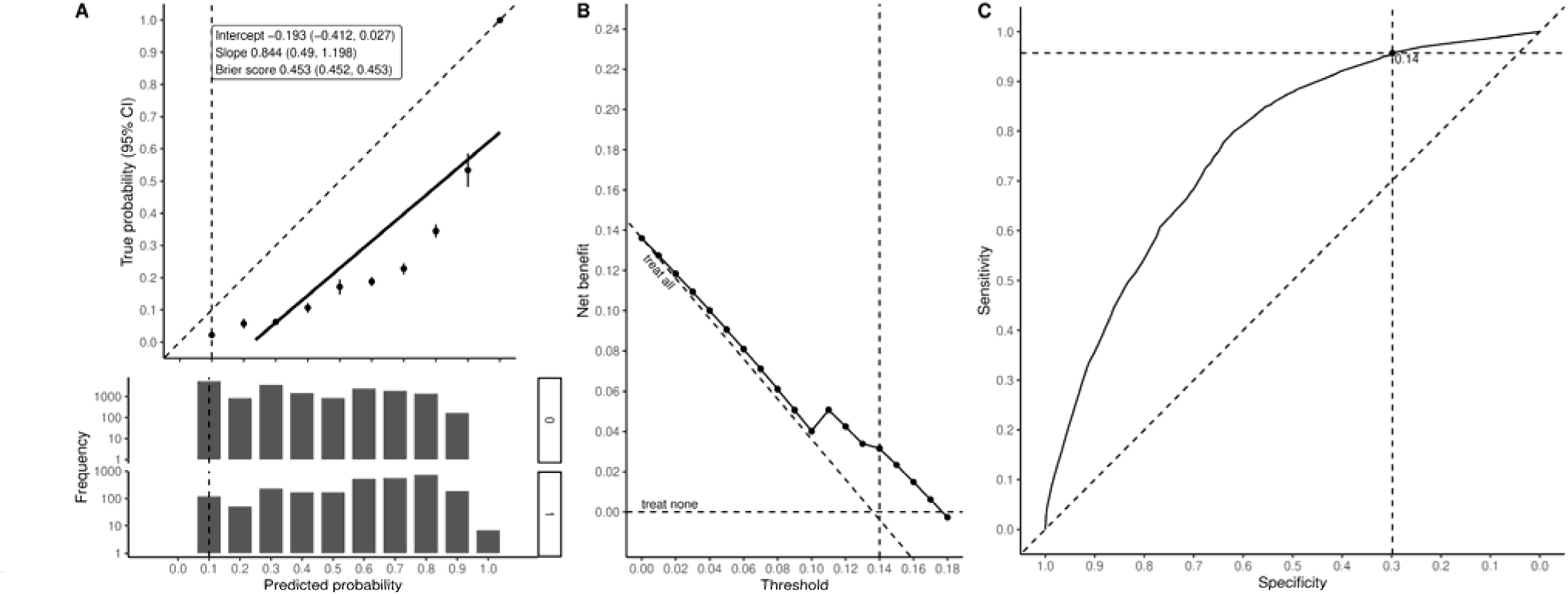
Evaluation of the best prediction model using the validation set: (A) calibration plot and the predicted probability distribution; (B) clinical utility by the decision curve; and (C) discrimination ability by the ROC curve. A regression line is fitted over the point estimates of the true probabilities on the calibration plot. The predicted probability distribution is also shown for either CA-UTI positive (=1) or negative (=0). The chosen threshold of 0.14 is shown by a dashed, vertical line on the calibration plot and decision curve. The corresponding sensitivity and specificity using the chosen threshold are also shown by the dashed, horizontal and vertical lines on the ROC curve. CA-UTI, catheter-associated UTI; CI, confidence interval; ROC, receiver operating characteristics; UTI, urinary tract infection.

No prediction model had a threshold with the expected net benefit that also led to ∼90% specificity. However, we optimized the best prediction model by prioritizing the expected net benefit and a reasonable tradeoff between sensitivity and specificity (Figure 2B), i.e., a threshold of 0.14. It characterized the best prediction model as one with ∼95% sensitivity (Figure 2C).

Using the test set with this threshold, we found that the best prediction model achieved 97.63% (95% CI: 97.57%, 97.69%) sensitivity, 20.95% (95% CI: 20.89%, 21.00%) specificity, 22.85% (95% CI: 22.79%, 22.92%) PPV, and 97.36% (95% CI: 97.29%, 97.42%) NPV.

We assessed the explainability of the best prediction model by the SHAP value. At the population level, a previous UTI diagnosis >2 times per six months or >3 times per year demonstrated the most extreme impact on the predicted positives (Figure 3A). Unlike this predictor, the impact of urological issues or a hospital stay ≥4 days was less extreme, but both predictors were more commonly positive. The impact of the latter predictor was also the highest on the predicted negatives with a hospital stay <4 days. Meanwhile, at the individual level, we identified two visually distinguished groups of clusters using four centers (*k*=4) (Figure S5): (1) clusters 1 and 2 and (2) clusters 3 and 4. All of the clusters covered true positives, false positives, and true negatives, but only clusters 1 and 2 covered false negatives, from which we randomly took a sample representing each. We show individual SHAP values for all samples (Figure S6). Unlike samples from clusters 3 and 4, those from clusters 1 and 2 showed two pairs of samples such that each pair shared the same predictor values, but one of each pair was falsely predicted. The four samples mentioned above covered both the predicted positives and negatives. To identify why the best prediction model made an error, we showed the values of unselected candidate predictors due to the limitations of our dataset (Figures 3B to 3E). Among them, more predictor values were respectively higher and lower for the true positive and negative samples. This finding indicated that the best model would have been more predictive if our dataset could provide those predictor values in a low-bias manner, i.e., neither perfect separation problem nor MNAR (before or after assigning their outliers as missing if any). The finding was confirmed at the population level by the lower intercept of the regression line (Figure 2A). Furthermore, the calibration plot showed that the less-extreme, predicted probabilities (i.e., ≥0.15 to <0.95) underestimated the true probabilities.

**Figure 3.**
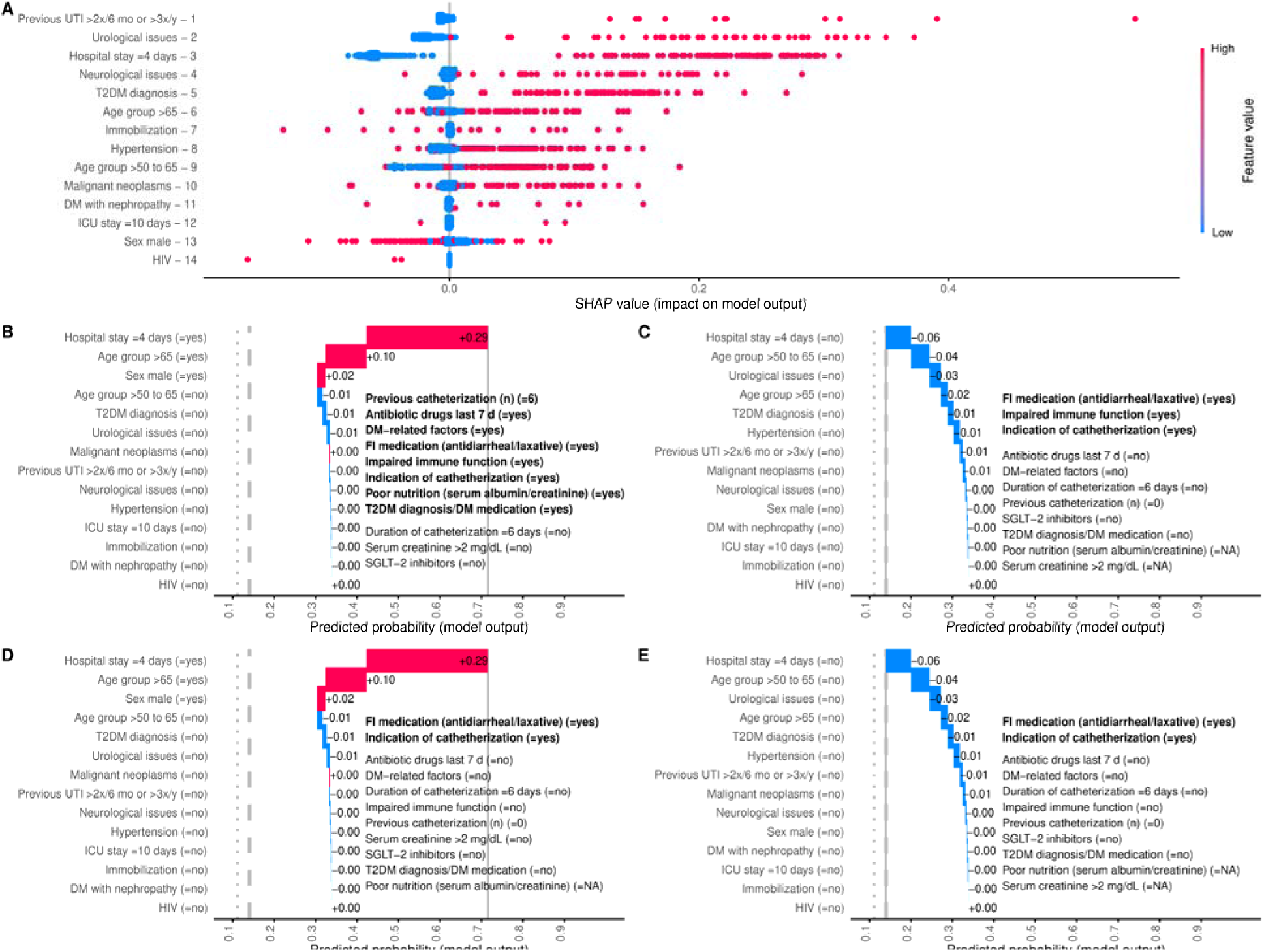
Explainability of the best prediction model at the population and individual levels: (A) beeswarm plot; (B) waterfall plot of a true positive from clusters 1 and 2; (C) waterfall plot of a false negative from clusters 1 and 2; (D) waterfall plot of a false positive from clusters 1 and 2; and (E) waterfall plot of a true negative from clusters 1 and 2. In panels B to E, the predicted probability of an individual starts from the average value among all individuals in the training set on the x-axis and from the least impactful predictor on the y-axis. SHAP values in panels B to E are shown at the end of the bars. The SHAP value may be zero due to rounding. The bar of a predictor with an absolute zero SHAP value is invisible. By adding SHAP values of the predictors, the end of the final bar reaches the predicted probability (solid line). If it is more than the threshold (dashed line), then the individual is predicted to be CA-UTI positive. As comparison, we also show the true probability (dotted line) and the values of candidate predictors that were unselected due to limitations of our dataset in panels B to E. The unselected candidate predictor with a higher value is in bold. See samples from other clusters in Figure S6. CA-UTI, catheter-associated UTI; DM, diabetes mellitus; FI, fecal incontinence; HIV, human immunodeficiency virus; ICU, intensive care unit; NA, not available; SGLT-2, sodium-glucose co-transporter-2; SHAP, the Shapley additive explanation; T2DM, type 2 DM; UTI, urinary tract infection.

## Discussion

### Finding summary

We developed and externally validated an RF model to predict CA-UTIs in advance. The predictors were: (1) age group >50 to 65 years old (no/yes), (2) age group >65 years old (no/yes), (3) ICU stay ≥10 days(no/yes), (4) hospital stay ≥4 days(no/yes), (5) sex male (no/yes), (6) T2DM diagnosis (no/yes), (7) immobilization (no/yes), (8) hypertension (no/yes), (9) DM with nephropathy (no/yes), (10) HIV (no/yes), (11) malignant neoplasms (no/yes), (12) urological issues (no/yes), (13) previous UTI diagnosis >2 times per six months or >3 times per year (no/yes), and (14) neurological issues. Our prediction model could detect 97.63% (95% CI: 97.57%, 97.69%) of CA-UTI positives. Among individuals that were predicted CA-UTI negatives, it was truly negatives by 97.36% (95% CI: 97.29%, 97.42%). Meanwhile, if an individual was predicted to be CA-UTI positive, it was truly positive by 22.85% (95% CI: 22.79%, 22.92%).

### Comparison to previous studies

Our model can predict CA-UTIs among hospitalized individuals receiving urinary catheterization. A prediction could also be made up to 48 h after catheter removal. Two previous studies developed prediction models for predicting CA-UTIs among ≥18-year-old individuals receiving urinary catheterization (indwelling), limited to those who had stayed in the ICU for ≥48 h due to neurologic conditions [4, 5].

In a single hospital, the first of the two previous studies developed (*n*=389; 4.37% CA-UTI positives) and internally validated (*n*=148; 4.37% CA-UTI positives) a prediction model [4]. It applied a logistic regression algorithm and used age group (<60/≥60 years old), admission diagnosis (brain tumor/brain injury/cerebral hemorrhage/aneurysm/epilepsy), ICU stay (<7/7 to 14/15 to 30/>30 days), and serum albumin (≥3.5/<3.5 g/dL). Unfortunately, the number of events in the training set (*n*=17) was too small for a regression algorithm, i.e., a minimum of 20 events per predictor [20]. The number of events in the validation set (*n*=4) was also too small, which was supposed to be at least 100 events [21].

In the same hospital and period of the data collection, the second study developed but did not validate their prediction model [5]. It applied a decision tree algorithm and used age group (<60/≥60 years old), admission diagnosis (brain tumor, brain injury, cerebral hemorrhage, or aneurysm/epilepsy), serum albumin (≥3.5/<3.5 g/dL), duration of catheterization (<7/≥7 days), sex (female/male), admission season (spring, autumn, or winter/summer), the Glasgow coma scale (>8/≤8), the number of antibiotic regimens (>2/≤2), and ventilator improvement (no/yes). The potential problem of sample size was noted, i.e., the number of events in the training set (*n*=21) which was less than 50 events per predictor, as required for a tree algorithm [20].

### Clinical implication

CA-UTIs represent a significant healthcare challenge due to their prevalence, impacts on patient outcomes, and contribution to antimicrobial resistance. Effective prevention, surveillance, and management strategies are crucial in mitigating their impacts. In addition, among other catheter-based strategies, a catheter with an antimicrobial inner surface is a potential substitute for conventional indwelling catheters [22, 23]. However, this recommendation lacks data to support its effectiveness in the general population [23]. A meta-analysis found that the efficacy of an antimicrobial catheter (i.e., silver-allow coated) for CA-UTIs was doubtful compared to that of a latex catheter, especially among either non-urology patients or those in institutions with a low prevalence of CA-UTIs [24]. Two clinical trials were conducted among non-urology patients. In the first trial, which enrolled neurosurgery patients, an ionic silver-impregnated catheter demonstrated significantly longer time to asymptomatic bacteriuria among patients with an indwelling catheter for a minimum of 24 h without postoperative antibiotics, compared to a silver-alloy, silicon Foley catheter but insignificant among the intention-to-treat population [25]. Meanwhile, the second trial enrolled urogynecology patients and found no significant difference in UTIs within six weeks of surgery between a silver alloy-impregnated suprapubic catheter and a standard latex one [26]. These findings implied that only individuals at high risk of a CA-UTI may benefit from an antimicrobial catheter. A risk prediction model such as we developed in our study is needed for this scenario.

There are two potential starting time points of clinical workflow into which our prediction model application may fit, i.e., when a clinician encounters (Figure 4): (1) a patient; (2) a patient with urinary catheterization. The first time point covers patients with urinary catheterization and those for which it is removed in ≤48 h. However, this application may require a policy integrated as a part of a hospital system, e.g., electronic health records (EHRs). The second time point is more practical for applying our prediction model, although it may uncover patients for which urine catheterization is removed in ≤48 h. This second approach can be a part of a checklist that ensures safe urine catheterization practice.

**Figure 4.**
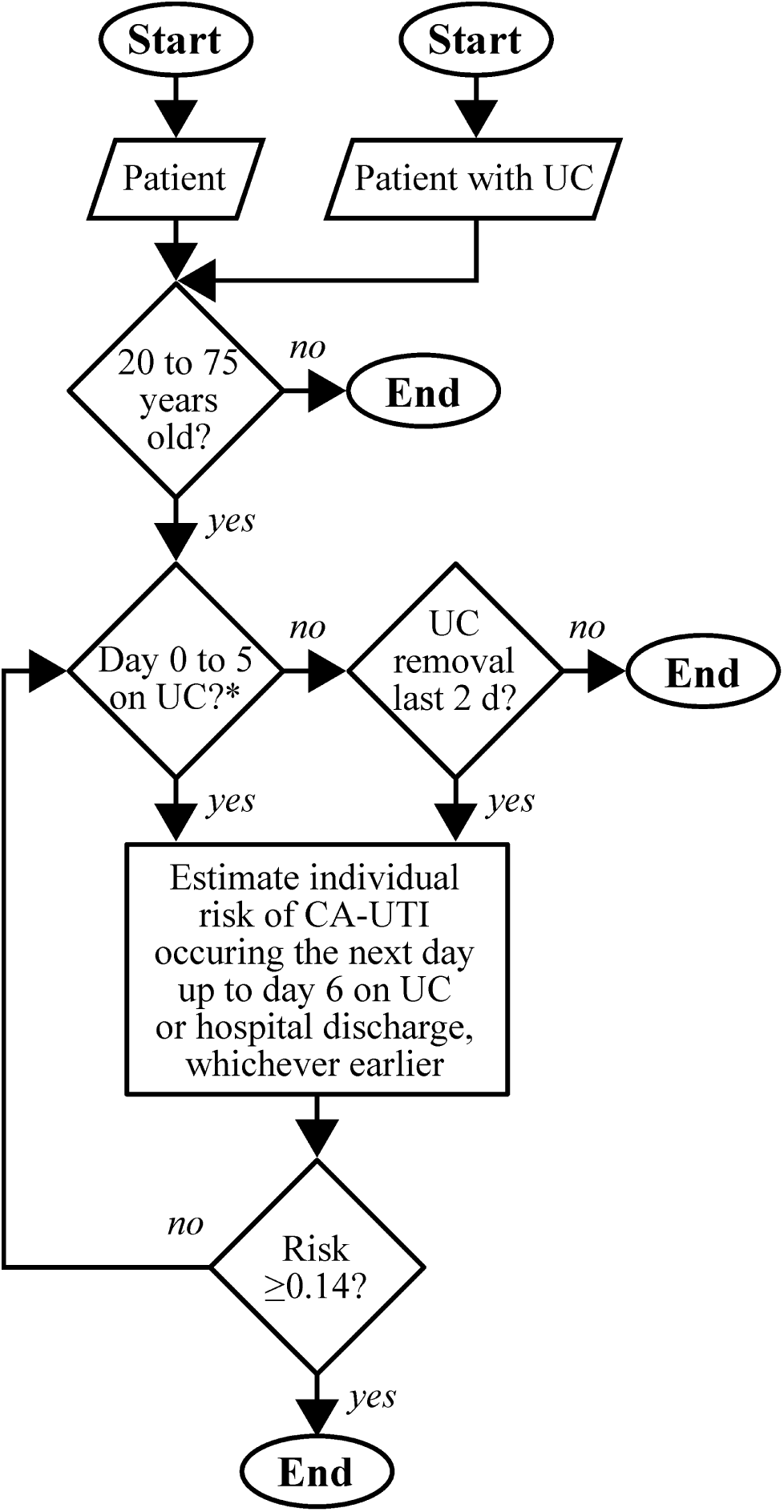
Proposed workflow for applying the prediction model. *, if the interval between this UC episode and the previous one is ≤1 day, then the day counted continues from the previous UC episode(s) until the interval >1 days; CA-UTI, catheter-associated urinary tract infection; UC, urine catheterization.

With the proposed workflow (Figure 4), clinicians can estimate the individual risks of CA-UTIs using our model. It can be embedded in an EHR system, for a clinical support system, and it is also provided by our web application for quick implementation (https://predme.app/en/cauti6d/). Furthermore, we provide a nomogram if a user cannot access the web application (Figure 5A). To use this nomogram, a user assesses all of the predictors from the first to the last order (Figures 5B and 5C) using these steps: (1) find a predictor with a positive value; (2) draw a line from the left-most-sided positive (=1) rectangle to the right-most-sided rectangle (either positive or negative); (3) match the predictor values to the data, upper and lower to the starting point of the line; (4) if they match, then draw a line from the lowest to the highest rectangles and find which panel this line ends up to estimate the risk (≥0.14 or <0.14); (5) if they do not match, then check the next right-hand-sided positive (=1) rectangle and repeat steps 4 to 5 until a rectangle of the next positive predictor is available downward; and (6) repeat step 1.

**Figure 5.**
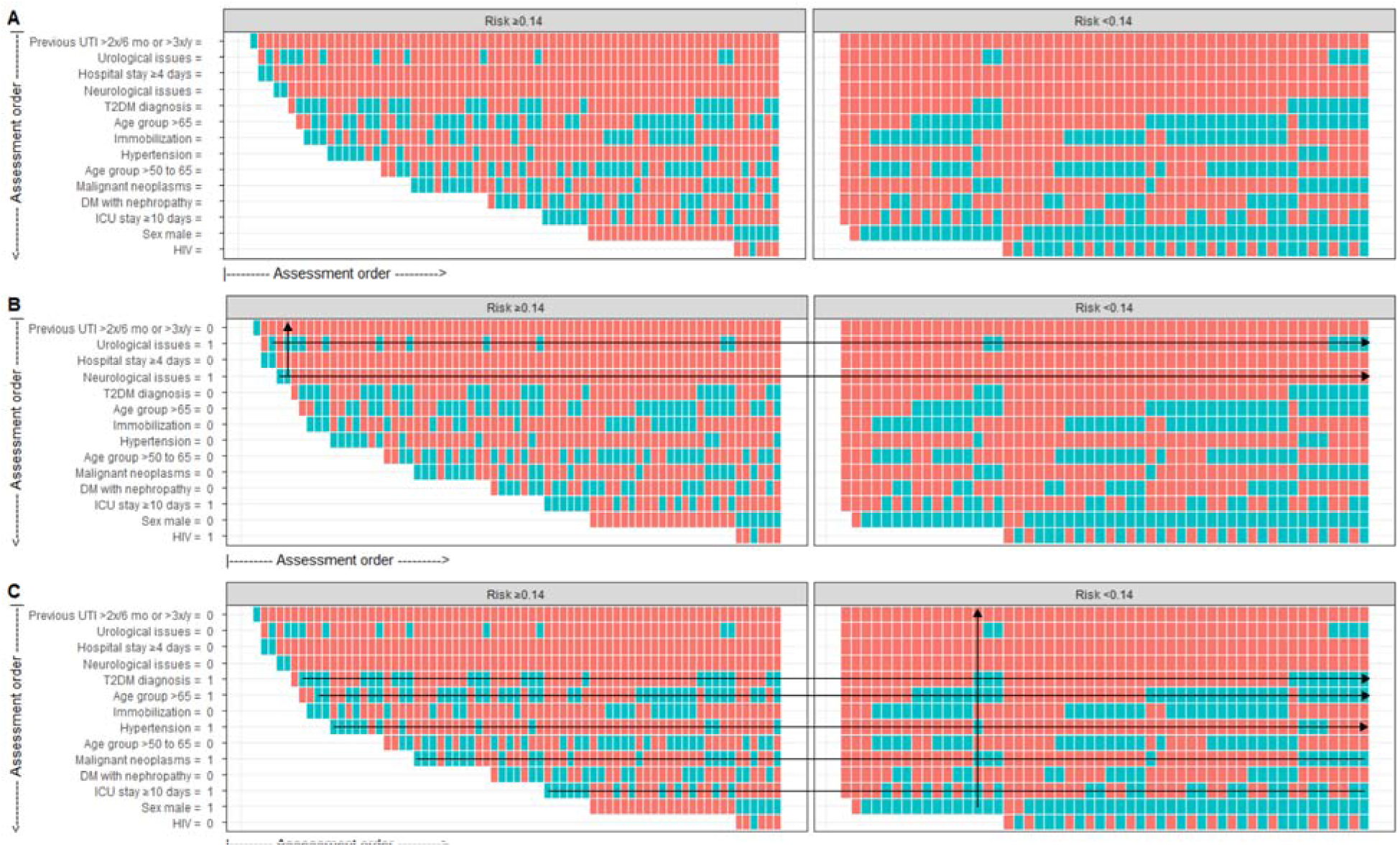
Nomogram: (A) empty; (B) predicted positive; (C) predicted negative. Cyan and red colors respectively indicate positive (=1) and negative (=0) values of each predictor. DM, diabetes mellitus; HIV, human immunodeficiency virus; ICU, intensive care unit; SGLT-2, sodium-glucose co-transporter-2; T2DM, type 2 DM; UTI, urinary tract infection.

If an individual’s risk of a CA-UTI is ≥0.14, then this workflow should not be used anymore for this episode of urinary catheterization. Any decisions after information of a positive prediction had not been available when our prediction model was developed. Such information can change the causal structure and our model’s predictive performance.

### Strengths and limitations

Our sample size was large for both model development and external validation. Furthermore, our prediction model achieved an acceptable predictive performance. In addition, our prediction model can be easily implemented. However, we need to address several limitations of our study. First, some variables could be biased in our database and thus might hamper the predictive performance of our model. Second, our model is also only capable of predicting a CA-UTI up to 6 days after the start of urinary catheterization before discharge. Third, our study cohort only consisted of an ethnic Chinese population, and data were collected in Taiwan. Since the epidemiology of CA-UTI varies globally, influenced by differences in healthcare practices and infection control policies, validation of our model in other countries should be considered. Finally, adherence to evidence-based guidelines is critical for reducing the incidence of CA-UTIs. Some healthcare-associated and behavioral variables, such as the compliance with the bundle care of urinary catheters and the drainage system, were not available in the database.

## Conclusions

A prediction model for CA-UTIs was developed and externally validated, using 14 predictors. Most CA-UTI positives could be detected, and most individuals were true negatives if they were predicted to be CA-UTI negatives. Therefore, our prediction model was clinically sufficient to support decision-making for preventing CA-UTIs among individuals which were predicted to be CA-UTI positives. Future studies are needed for prospective evaluation of our prediction by EHR integration, web application, or a nomogram. Individualized treatment after knowing the prediction should be developed and tested. Eventually, we also need to evaluate the impacts of the prediction model with or without specific treatments on patient outcomes.

## Supporting information

TRIPOD checklist

## Data Availability

The data that support the findings of this study are available from the Clinical Data Center, Office of Data Science, Taipei Medical University, Taiwan, but restrictions apply to the availability of these data, which were used under license for the current study (access approval no.: A202206008), and so are not publicly available. Data are however available from the authors upon reasonable request and with permission of the Clinical Data Center. To get this permission, one need to request an access from the Clinical Data Center (https://ods.tmu.edu.tw/). We shared the programming codes for all analyses in this study (https://github.com/herdiantrisufriyana/colab_uti), including predictive modeling.

## Acknowledgements

The authors acknowledge the computational and technical support of the Clinical Data Center, Office of Data Science, Taipei Medical University, Taiwan. This work was supported by: (1) the Postdoctoral Accompanies Research Project from the National Science and Technology Council in Taiwan [grant no. NSTC111-2811-E-038-003-MY2] to HS; (2) the Ministry of Science and Technology in Taiwan [grant nos. MOST110-2628-E-038-001 and MOST111-2628-E-038-001-MY2] to ECYS; and (3) the Higher Education Sprout Project from the Ministry of Education in Taiwan [grant nos. DP2-111-21121-01-A-05 and DP2-TMU-112-A-13] to ECYS. These funding bodies had no role in the study design; in the collection, analysis, and interpretation of the data; in the writing of the report; or in the decision to submit the article for publication. The authors declare that they have no competing interests.

## CRediT authorship contribution statement

**HS:** Conceptualization, Methodology, Software, Validation, Formal analysis, Writing – original draft, Visualization, Funding acquisition. **CC:** Conceptualization, Methodology, Validation, Investigation, Data curation, Writing – review & editing, Project administration. **HSC:** Conceptualization, Methodology, Writing – review & editing. **PS:** Conceptualization, Methodology, Writing – review & editing. **PYY:** Conceptualization, Methodology, Validation, Investigation, Data curation, Writing – review & editing, Project administration. **JHK:** Conceptualization, Methodology, Resources, Writing – review & editing, Supervision. **ECYS:** Conceptualization, Methodology, Resources, Writing – review & editing, Supervision, Funding acquisition. All authors have read and approved the manuscript and agreed to be accountable for all aspects of the work in ensuring that questions related to the accuracy or integrity of any part of the work are appropriately investigated and resolved.

