## Supplementary Materials for "Estimating individual risk of catheter-associated urinary tract infections using explainable artificial intelligence on clinical data"

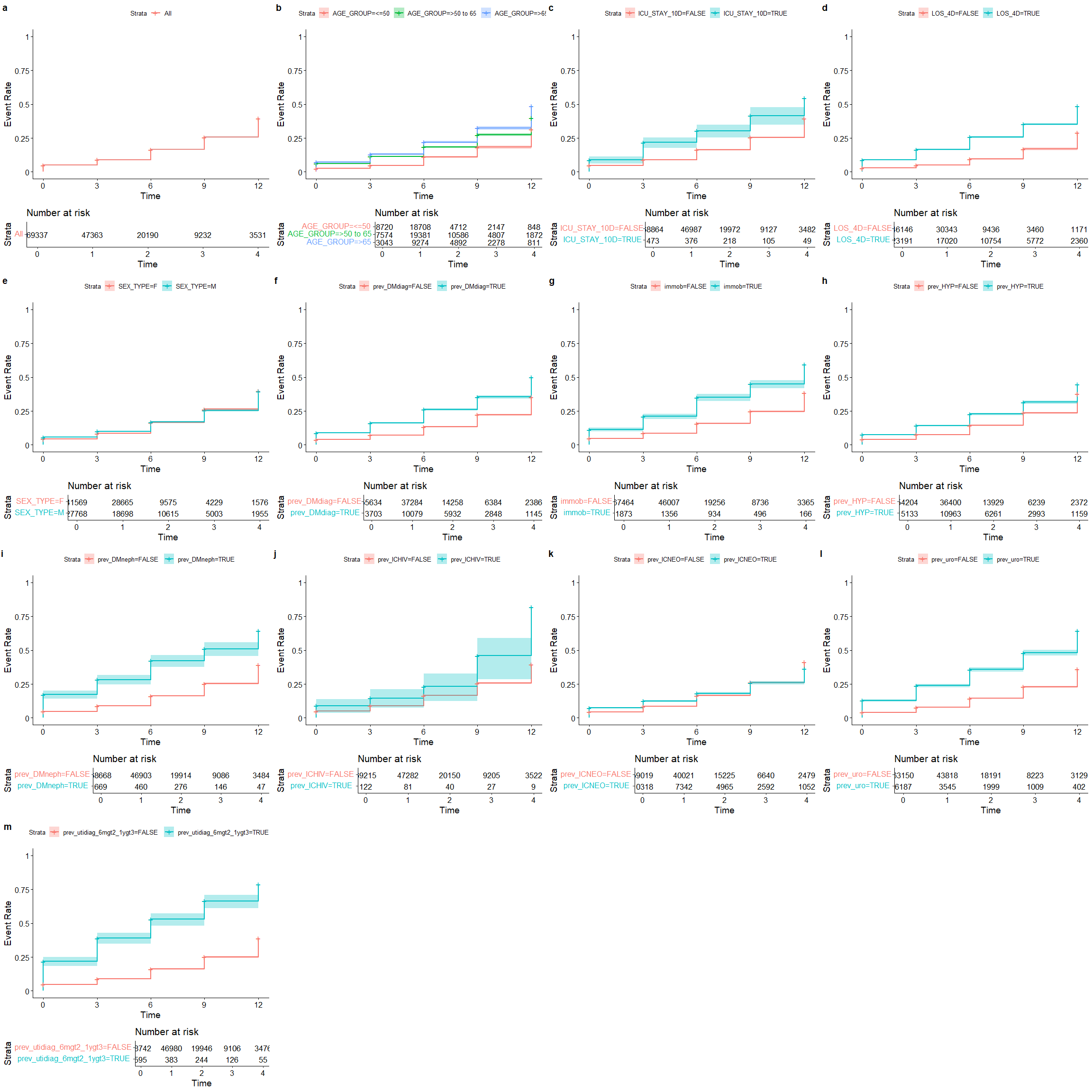

Figure S1. Selected span and range for outcome onset (A) and KM plot of univariate regression analyses for all the selected candidate predictors (B to M). Each time in x-axis refers to a day. AGE_GROUP, age group >50 to 65 and >65 years old; DM, diabetes mellitus; HR, hazard ratio; HIV, human immunodeficiency virus; ICU_STAY_10D, ICU stay ≥10 days; ICU, intensive care unit; immob, immobilization; LOS_4D, hospital stay ≥4 days; prev_DMdiag, T2DM diagnosis; prev_DMneph, DM with nephropathy; prev_HYP, hypertension; prev_ICHIV, HIV; prev_ICNEO, malignant neoplasms; prev_uro, urological issues; prev_utidiag_6mgt2_1ygt3, previous UTI diagnosis >2 times per six months or >3 times per year; SEX_TYPE, sex male; T2DM, type 2 DM; UTI, urinary tract infection.

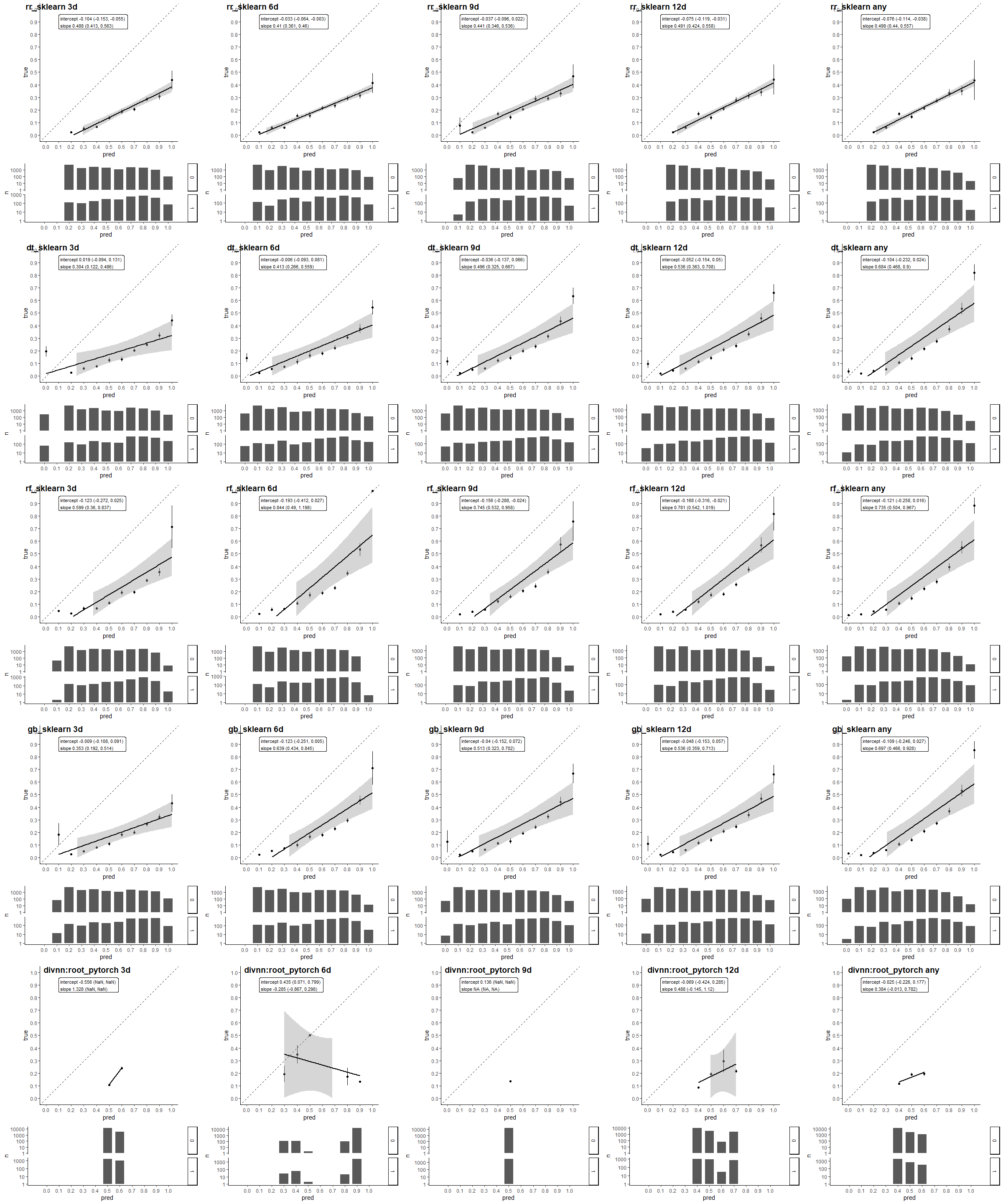

Figure S2. Calibration of the prediction models using the validation set. The intercept and slope are stated in a point and interval estimates (i.e., 95% CI). any, predict CA-UTI without a specific onset; CA-UTI, catheter-associated UTI; CI, confidence interval; d, predict CA-UTI with a specific onset day (1 to 3, 6, 9, and 12); dt, decision tree; divnn:root, deep-insight visible neural network (root ontology prediction); rf, random forest; rr, ridge regression; gb, extreme gradient boosting; UTI, urinary tract infection.

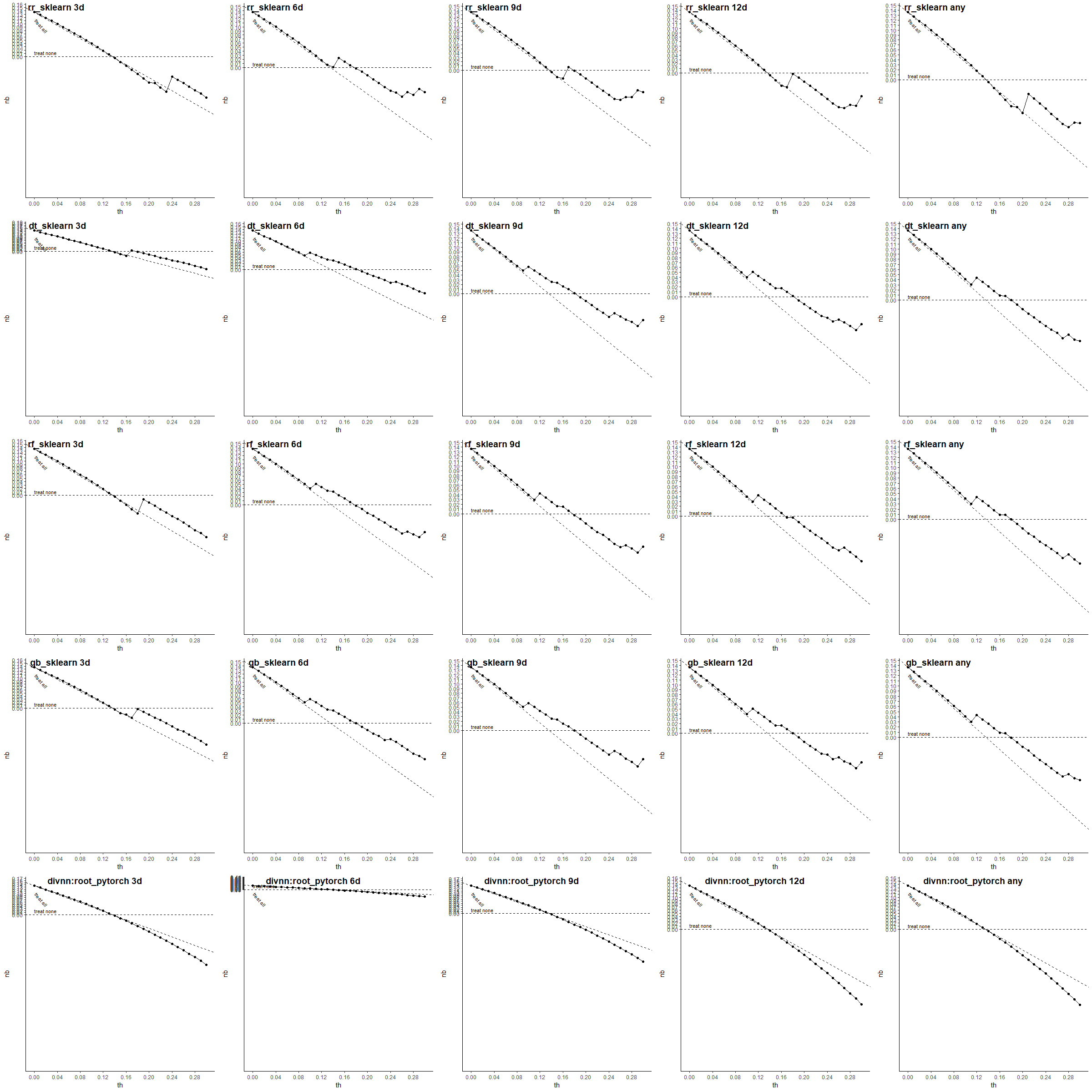

Figure S3. Decision curve analysis of the prediction models using the validation set. any, predict CA-UTI without a specific onset; CA-UTI, catheter-associated UTI; d, predict CA-UTI with a specific onset day (1 to 3, 6, 9, and 12); dt, decision tree; divnn:root, deep-insight visible neural network (root ontology prediction); rf, random forest; rr, ridge regression; gb, extreme gradient boosting; UTI, urinary tract infection.

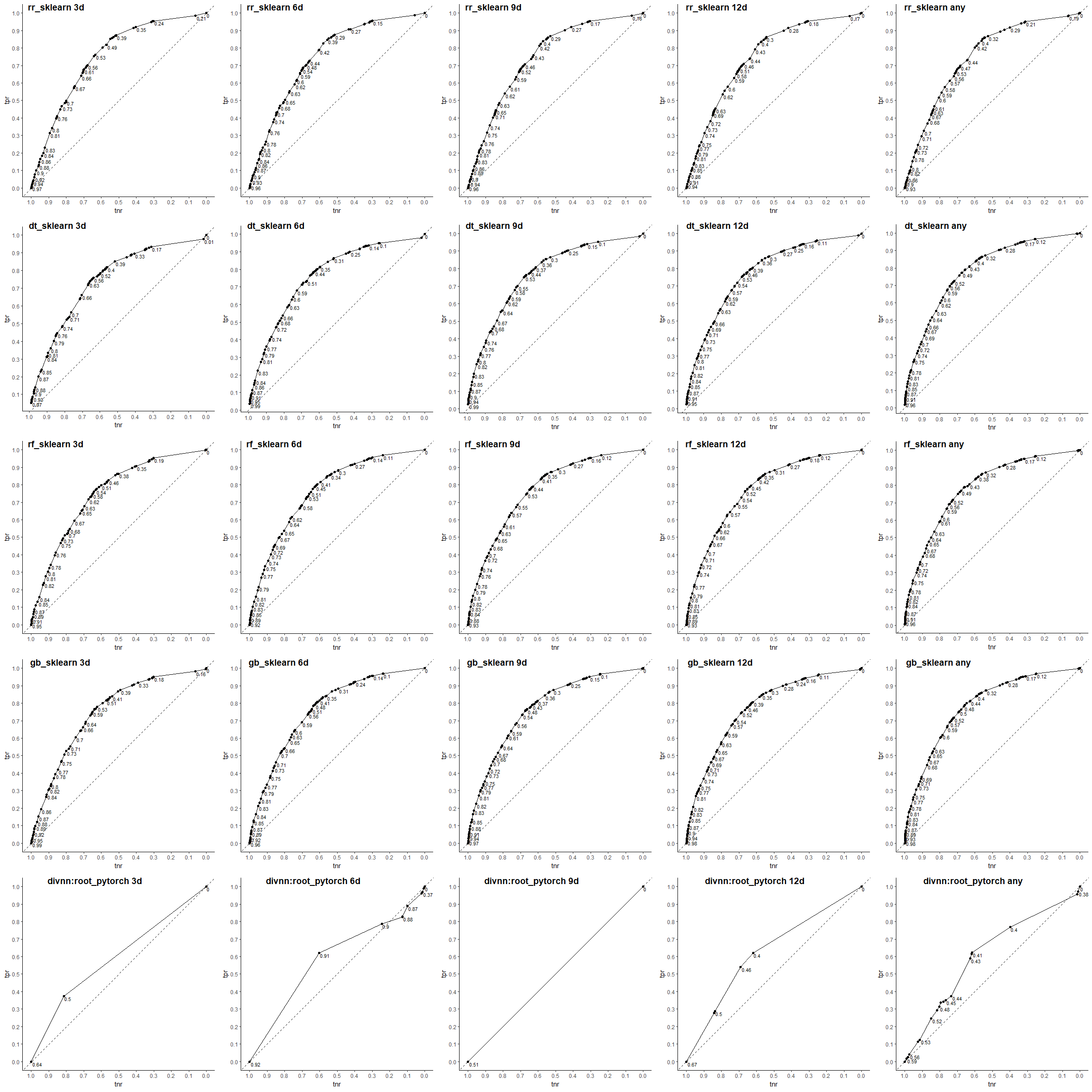

Figure S4. ROC of the prediction models using the validation set. Each point on the curve is labeled by a threshold. any, predict CA-UTI without a specific onset; CA-UTI, catheter-associated UTI; d, predict CA-UTI with a specific onset day (1 to 3, 6, 9, and 12); dt, decision tree; divnn:root, deep-insight visible neural network (root ontology prediction); rf, random forest; ROC, receiver operating characteristics; rr, ridge regression; gb, extreme gradient boosting; UTI, urinary tract infection.

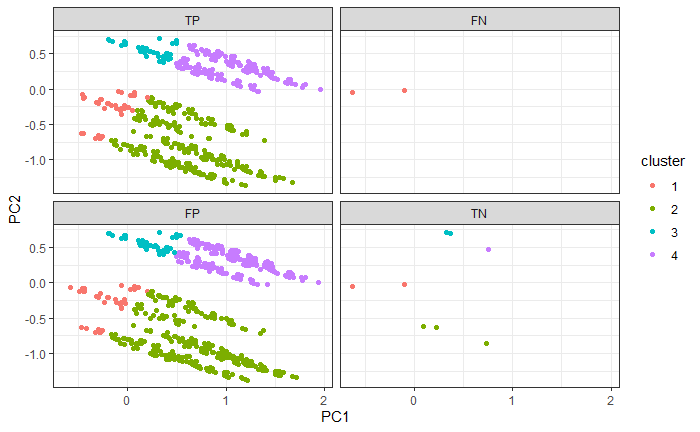

Figure S5. Visually distinguished groups of clusters using four centers (*k*=4). FN, false negative; FP, false positive; PC, principal component; TN, true negative; TP, true positive.

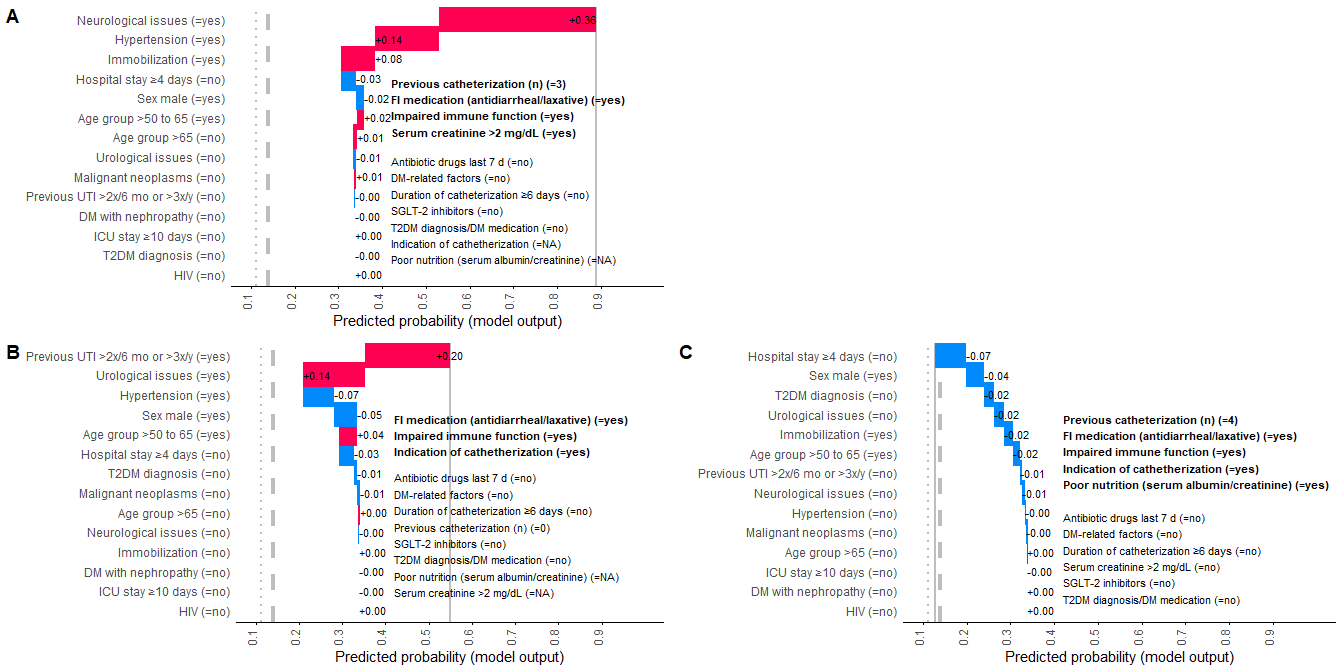

Figure S6. Explainability of the best prediction model at individual level from samples from other clusters: (A) waterfall plot of a true positive from clusters 3 and 4; (B) waterfall plot of a false positive from clusters 3 and 4; and (C) waterfall plot of a true negative from clusters 3 and 4. In panels A to C, the predicted probability of individual starts from the average value among all individuals in the training set by x-axis and from the least impactful predictor by y-axis. SHAP values in panels A to C are shown at the end of the bars. The shown SHAP value may be zero due to rounding. The bar of a predictor with absolute zero SHAP value is unseen. By adding SHAP values of the predictors, the end of the final bar reaches the predicted probability (solid line). If it is more than the threshold (dashed line), then the individual is predicted CA-UTI positive. As comparison, we also show the true probability (dotted line) and the values of candidate predictors that were unselected due to limitations of our dataset in panels A to C. The unselected candidate predictor with a higher value is bold. CA-UTI, catheter-associated UTI; DM, diabetes mellitus; FI, fecal incontinence; HIV, human immunodeficiency virus; ICU, intensive care unit; NA, not available; SGLT-2, sodium-glucose co-transporter-2; SHAP, the Shapley additive explanation; T2DM, type 2 DM; UTI, urinary tract infection.

Table S1. Hyperparameter grid of the machine learning algorithms.

| Algorithm | Hyperparameter |
| --- | --- |
| Ridge regression | C: [0.01,0.1,1,10] |
| Decision tree | max_depth: [None,10,20,30]  min_samples_split: [2,5,10] |
| Random forest | n_estimators: [10,50,100]  max_depth: [None,10,20]  min_samples_split: [2,5,10]} |
| Extreme gradient boosting | n_estimators: [50,100,200]  learning_rate: [0.01,0.1,1]  max_depth: [3,4,5]  subsample: [0.8,1]  colsample_bytree: [0.8,1] |
| Deep-insight visible neural network | l2_norm:[0.01,0.1,1,10] |

Table S2. Candidate predictors and their indicators.

| Candidate predictor | Indicator |  |
| --- | --- | --- |
| 1. Age group | 1. Age (years) |  |
| 2. ICU stay >=10 days | 2. ICU stay (days) |  |
| 3. Duration of catheterization >=6 | 3. Duration of catheterization (days) |  |
| 4. Hospital stay >=4 days | 4. Hospital stay (days) |  |
| 5. Previous catheterization (n) | 5. Previous catheterization (n) |  |
| 6. Sex (F/M) | 6. Sex (F/M) |  |
| 7. T2DM diagnosis/DM medication | 7. T2DM diagnosis |  |
|  | 8. DM medication |  |
| 8. Immobilization (protective restraint) | 9. Immobilization (protective restraint) |  |
| 9. Indication of catheterization | 10. Indication of catheterization |  |
| 10. DM-related factors | 11. Hypertension diagnosis |  |
|  | 12. Exogenous insulin |  |
|  | 13. BMI >30 | 13.1. BMI (kg/m^2^) |
|  | 14. DM with nephropathy |  |
| 11. Poor nutrition (serum albumin/creatinine) | 15. Serum albumin <3.5 g/dL | 15.1. Serum albumin (g/dL) |
|  | 16. Serum creatinine <0.5 mg/dL & eGFR >=105 mL/min/1.73 m^2^ | 16.1. Serum creatinine (mg/dL) |
|  |  | 16.2. eGFR (mL/min/1.73 m^2^) |
| 12. FI medication (antidiarrheal/laxative) | 17. Antidiarrheal drugs |  |
|  | 18. Laxative drugs |  |
| 13. Impaired immune function | 19. Corticosteroid drugs |  |
|  | 20. HIV |  |
|  | 21. Malignant neoplasms |  |
| 14. Serum creatinine >2 mg/dL | 22. Serum creatinine >2 mg/dL |  |
| 15. SGLT-2 inhibitors | 23. SGLT-2 inhibitors |  |
| 16. Urological issues (double-J/diagnosis/urodynamics) | 24. Double-J ureteral stent insertion |  |
|  | 25. Urological diagnosis |  |
|  | 26. Urodynamic examination |  |
| 17. Previous UTI diagnosis >2x/6 mo or >3/1 y | 27. Previous UTI diagnosis >2x/6 mo | 27.1. Previous UTI diagnosis last 6 mo (n) |
|  | 28. Previous UTI diagnosis >3/1 y | 28.1. Previous UTI diagnosis last 1 y (n) |
| 18. Neurological issues (diagnosis) | 29. Neurological issues (diagnosis) |  |
| 19. Antibiotic drugs last 7 d | 30. Antibiotic drugs last 7 d |  |

BMI, body mass index; DM, diabetes mellitus; eGFR, estimated glomerular filtration rate; F, female; HIV, human immunodeficiency virus; ICU, intensive care unit; M, male; SGLT, sodium-glucose co-transporter; T2DM, type 2 DM; UTI, urinary tract infection.

Table S3. Baseline characteristics of validation and test sets.

| Variable | | CA-UTI | | | | |
| --- | --- | --- | --- | --- | --- | --- |
|  |  | Validation set | | | Test set | |
|  |  | No (*n*=17,497) | Yes (*n*=2756) | No (*n*=16,986) | | Yes (*n*=4074) |
| Age group | ≤50 years old, % (*n*) | 42.25 (7392) | 21.63 (596) | 37.74 (6410) | | 22.41 (913) |
|  | >50 to 65 years old, % (*n*) | 39.55 (6920) | 50.69 (1397) | 40.87 (6943) | | 51.4 (2094) |
|  | >65 years old, % (*n*) | 18.2 (3185) | 27.69 (763) | 21.39 (3633) | | 26.19 (1067) |
| ICU stay ≥10 days | No, % (*n*) | 99.33 (17,380) | 98.33 (2710) | 98.48 (16,727) | | 97.13 (3957) |
|  | Yes, % (*n*) | 0.67 (117) | 1.67 (46) | 1.52 (259) | | 2.87 (117) |
| Hospital stay ≥4 days | No, % (*n*) | 65.93 (11,536) | 31.42 (866) | 42.84 (7277) | | 27.54 (1122) |
|  | Yes, % (*n*) | 34.07 (5961) | 68.58 (1890) | 57.16 (9709) | | 72.46 (2952) |
| Sex | Female, % (*n*) | 58.88 (10,302) | 52.07 (1435) | 54.05 (9181) | | 55.18 (2248) |
|  | Male, % (*n*) | 41.12 (7195) | 47.93 (1321) | 45.95 (7805) | | 44.82 (1826) |
| T2DM diagnosis | No, % (*n*) | 81.38 (14,239) | 62.55 (1724) | 73.43 (12,473) | | 57.44 (2340) |
|  | Yes, % (*n*) | 18.6 (3255) | 37.45 (1032) | 26.55 (4510) | | 42.56 (1734) |
| Immobilization | No, % (*n*) | 97.34 (17,031) | 91.69 (2527) | 87.16 (14,805) | | 83.68 (3409) |
|  | Yes, % (*n*) | 2.66 (466) | 8.31 (229) | 12.84 (2181) | | 16.32 (665) |
| Hypertension | No, % (*n*) | 78.14 (13,673) | 64.55 (1779) | 72.36 (12,291) | | 67.72 (2759) |
|  | Yes, % (*n*) | 21.84 (3821) | 35.45 (977) | 27.62 (4692) | | 32.28 (1315) |
| DM with nephropathy | No, % (*n*) | 99.09 (17,338) | 97.35 (2683) | 98.54 (16,738) | | 96.61 (3936) |
|  | Yes, % (*n*) | 0.89 (156) | 2.65 (73) | 1.44 (245) | | 3.39 (138) |
| HIV | No, % (*n*) | 99.87 (17,474) | 99.64 (2746) | 99.89 (16,968) | | 99.85 (4068) |
|  | Yes, % (*n*) | 0.11 (20) | 0.36 (10) | 0.09 (15) | | 0.15 (6) |
| Malignant neoplasms | No, % (*n*) | 83.97 (14,693) | 76.71 (2114) | 84.09 (14,283) | | 83.55 (3404) |
|  | Yes, % (*n*) | 16.01 (2801) | 23.29 (642) | 15.9 (2700) | | 16.45 (670) |
| Urological issues | No, % (*n*) | 92.54 (16,191) | 80.08 (2207) | 81.18 (13,790) | | 68.92 (2808) |
|  | Yes, % (*n*) | 7.45 (1303) | 19.92 (549) | 18.8 (3193) | | 31.08 (1266) |
| Previous UTI diagnosis >2 times per six months or >3 times per year | No, % (*n*) | 97.79 (17,110) | 95.46 (2631) | 95.52 (16,225) | | 91.04 (3709) |
|  | Yes, % (*n*) | 0.52 (91) | 2.69 (74) | 2.45 (417) | | 6.77 (276) |
| Neurological issues | No, % (*n*) | 92.93 (16,260) | 81.64 (2250) | 79.54 (13,511) | | 77.1 (3141) |
|  | Yes, % (*n*) | 7.05 (1234) | 18.36 (506) | 20.44 (3472) | | 22.9 (933) |

Total percentage (%) and number (*n*) may not be respectively 100% and the same with the number of an outcome category, because it includes data with missing values. CA-UTI, catheter-associated UTI; DM, diabetes mellitus; HIV, human immunodeficiency virus; ICU, intensive care unit; T2DM, type 2 DM; UTI, urinary tract infection.

Table S4. The covariates for the adjustment.

| Candidate predictor | | Covariate code | | | | | | | | | | | | |
| --- | --- | --- | --- | --- | --- | --- | --- | --- | --- | --- | --- | --- | --- | --- |
| Name | Code | 1 | 2 | 3 | 4 | 5 | 6 | 7 | 8 | 9 | 10 | 11 | 12 | 13 |
| Age group >50 to 65 years old | 1 | 0 | 1 | 1 | 1 | 1 | 1 | 1 | 1 | 1 | 1 | 1 | 1 | 1 |
| Age group >65 years old | 1 | 0 | 1 | 1 | 1 | 1 | 1 | 1 | 1 | 1 | 1 | 1 | 1 | 1 |
| ICU stay ≥10 days | 2 | 1 | 0 | 0 | 1 | 1 | 0 | 1 | 0 | 0 | 0 | 0 | 0 | 1 |
| Hospital stay ≥4 days | 3 | 1 | 0 | 0 | 1 | 1 | 1 | 1 | 1 | 1 | 1 | 1 | 1 | 1 |
| Sex male | 4 | 1 | 1 | 1 | 0 | 1 | 1 | 1 | 0 | 1 | 1 | 1 | 1 | 1 |
| T2DM diagnosis | 5 | 1 | 1 | 1 | 1 | 0 | 1 | 1 | 0 | 1 | 1 | 1 | 1 | 1 |
| Immobilization | 6 | 1 | 0 | 1 | 1 | 1 | 0 | 1 | 1 | 0 | 1 | 1 | 1 | 1 |
| Hypertension | 7 | 1 | 1 | 1 | 1 | 1 | 1 | 0 | 1 | 1 | 1 | 1 | 1 | 1 |
| DM with nephropathy | 8 | 1 | 0 | 1 | 0 | 0 | 1 | 1 | 0 | 0 | 1 | 1 | 1 | 1 |
| HIV | 9 | 1 | 0 | 1 | 1 | 1 | 0 | 1 | 0 | 0 | 0 | 0 | 0 | 1 |
| Malignant neoplasms | 10 | 1 | 0 | 1 | 1 | 1 | 1 | 1 | 1 | 0 | 0 | 1 | 1 | 1 |
| Urological issues | 11 | 1 | 0 | 1 | 1 | 1 | 1 | 1 | 1 | 0 | 1 | 0 | 1 | 1 |
| Previous UTI diagnosis >2 times per six months or >3 times per year | 12 | 1 | 0 | 1 | 1 | 1 | 1 | 1 | 1 | 0 | 1 | 1 | 0 | 1 |
| Neurological issues | 13 | 1 | 1 | 1 | 1 | 1 | 1 | 1 | 1 | 1 | 1 | 1 | 1 | 0 |

Codes for candidate predictor and covariate columns are the same. A covariate is assigned to 1 if it was included in the adjustment; otherwise, it is assigned to 0. CA-UTI, catheter-associated UTI; CI, confidence interval; DM, diabetes mellitus; HR, hazard ratio; HIV, human immunodeficiency virus; ICU, intensive care unit; T2DM, type 2 DM; UTI, urinary tract infection.

Table S5. AUC-ROC of the prediction models using the validation set.

| Algorithm | AUC-ROC (95% CI) | | | |  |
| --- | --- | --- | --- | --- | --- |
|  | Predict CA-UTI with a specific onset (day) | | | | Predict CA-UTI without a specific onset |
|  | 1 to 3 | 1 to 6 | 1 to 9 | 1 to 12 |  |
| Ridge regression | 0.749  (0.747, 0.751) | 0.75  (0.748, 0.751) | 0.752  (0.75, 0.754) | 0.752  (0.75, 0.754) | 0.752  (0.75, 0.754) |
| Decision tree | 0.74  (0.738, 0.742) | 0.761  (0.759, 0.763) | 0.771  (0.769, 0.773) | 0.776  (0.774, 0.778) | 0.788  (0.786, 0.789) |
| Random forest | 0.751  (0.749, 0.753) | 0.768  (0.766, 0.77) | 0.773  (0.772, 0.775) | 0.776  (0.774, 0.778) | 0.788  (0.786, 0.79) |
| Extreme gradient boosting | 0.749  (0.747, 0.752) | 0.768  (0.766, 0.769) | 0.774  (0.772, 0.776) | 0.776  (0.775, 0.778) | 0.787  (0.785, 0.789) |
| Deep-insight visible neural network | 0.595  (0.593, 0.597) | 0.588  (0.587, 0.59) | 0.5  (0.5, 0.5) | 0.628  (0.626, 0.629) | 0.611  (0.608, 0.613) |

AUC, area under curve; CA-UTI, catheter-associated UTI; CI, confidence interval; ROC, receiver operating characteristics; UTI, urinary tract infection.
